# Higher platelet counts are associated with migraine risk across three cohorts among more than 21,000 cases

**DOI:** 10.64898/2026.11.09.26367616

**Authors:** Emily M. Sjodin, Daniel I. Chasman, Ming-Huei Chen, Pamela M. Rist, Andrew D. Johnson

**Author notes:** Corresponding author: Andrew D. Johnson, 73 Mt. Wayte Ave., Suite #2, Framingham, MA 01702.

## Abstract

**Objective:** We assessed potential associations between migraine and complete blood cell count (CBC) data in three large population-based studies.

**Background:** Platelets are thought to be involved in migraine pathology through increased activation during an attack. Migraine attacks are often treated with NSAIDs that inhibit platelets, or triptans that target serotonin receptors, with serotonin being most highly expressed in circulation within platelet granules. However, the exact role of platelets in migraine remains unclear.

**Methods:** The UK BioBank (UKBB) has CBC data available from baseline blood samples and migraine diagnoses were obtained through self-report and electronic health records. CBC and migraine data from the 1999-2000, 2001-2002, and 2003-2004 National Health and Nutrition Examination Survey (NHANES) study were obtained during single exam visits. CBCs were obtained from Framingham Heart Study (FHS) participants. Migraine history was assessed through clinical diagnostic impressions.

The UKBB sample (18,919 cases (4,580 incident); 448,339 controls) was the primary discovery cohort for CBCs, with replication meta-analysis done in NHANES (1,887 cases; 7,242 controls) and FHS (395 cases (229 incident); 3,172 controls). Our primary model adjusted for age and sex, with secondary analysis adjusting for additional factors. Sex-stratified, sex-interaction and ancestry-stratified (UKBB only) analyses were also performed.

**Results:** Migraine was associated with platelet count (PLT) in the UKBB (OR=1.058, 95% CI [1.043,1.073], p=2.09E-14) and was replicated in the NHANES-FHS meta-analysis (OR=1.074, 95% CI [1.025,1.126], p=0.003). PLT also drove multiple significant signals of PLT-involved CBC ratios. While neutrophil counts and hemoglobin were associated with migraine in UKBB, these did not replicate in FHS-NHANES meta-analysis.

**Conclusion:** Higher platelet count is associated with an increased likelihood of migraine. Further studies with longitudinal assessments of CBCs and migraine attacks are needed to further understand the role of platelets in the onset of migraine attacks.

## Introduction

Platelets are hypothesized to play an important role in migraine pathology, as migraine has a prothrombotic phenotype^[1]^. The potential linkage may be via a combination of endothelial dysfunction, leukocyte-platelet and leukocyte-endothelial interactions, and altered platelet activation^[2],[3]^. Additionally, serotonin is thought to play a role in migraine-platelet pathology as platelets share structural similarities to serotoninergic neurons^[4]^. Serotonin helps sensitize sensory neurons^[1]^ and is stored in the dense granules within platelets, representing the greatest pool of serotonin in the bloodstream. Therefore, changes to platelets might reflect changes to serotonergic pathways and function^[2]^. The platelets of individuals who experience migraine attacks might also have impaired serotonin metabolism^[1]^.

Blood cell counts, and ratios, have also been examined in studies. This includes reports of differences especially in platelet-platelet^[1],[5]^ and platelet-leukocyte aggregates^[1],[3],[5],[6]^, although some findings were specific to migraine attacks with or without aura^[7]^. The neutrophil-monocyte ratio^[8]^, neutrophil-leukocyte ratio^[6],[8]^, neutrophil-monocyte ratio^[8]^, and the monocyte-leukocyte ratio^[6]^ were also reported to be increased among those with migraine. However, a contrary paper found decreased platelet-leukocyte ratio among individuals with migraine^[8]^. Platelet count was investigated also with varying results, including no significant differences^[1],[9],[10]^, trends in several studies^[10^ ^11]^ and reports of higher platelet counts in several studies of chronic headache types^[12^ ^13]^. Recently a single biobank phenome study reported increased platelet count associated with migraine risk overall, and in women, though this study did not focus mainly on cell counts and did not include a replication design^[14].^ Platelet size ^[1]^, lymphocyte, or granulocyte counts were not previously associated with migraine^[10]^.

However, a major limitation to most prior work is relatively modest sample sizes and a lack of replication samples. Thus, most were underpowered to detect small or moderate associations between cell counts, cell count ratios, and migraine. Based on the previous literature, we hypothesized that platelet cell counts and cell ratios indicative of altered immune function are associated with migraine. We explored these hypotheses in larger cohorts and biobanks than prior studies, including a replication design.

## Methods

### UK Biobank

#### Participant Selection

The UK Biobank collected health data from over 500,000 people, including blood samples and lifestyle information^[15]^. Cell counts were measured at baseline. Migraine outcome was defined by self-report and ICD9 (3460, 3461, 3462, 3468, 3469) and ICD10 (G43.0, G43.1, G43.2, G43.3, G43.8, G43.9) codes at baseline and during follow-up. Incident cases had the same ICD codes with an occurrence later than the baseline date, or if the self-reported age at occurrence was older than baseline age. We assessed mean platelet volume (MPV), platelet count (PLT), lymphocyte (LYM), monocyte (MON), neutrophil (NEU), hemoglobin (HGB), red blood cell distribution width (RDW), white blood cell count (WBC) and nine cell count ratios constructed among these eight cell count traits. The cell count ratios (MPV.PLT, PLT.LYM, PLT.MON, PLT.NEU, HGB.RDW, RDW.PLT, NEU.LYM, MON.LYM, WBC.MPV) were selected based on biomarker studies. In ratio names (*e.g.* MPV.PLT), the first cell count (MPV) was the numerator, and the second cell count (PLT) was the denominator. Log transformation was applied to the derived cell count ratios^[16]^. Samples with NEU=0, LYM=0, without genetic data for inferring ancestry, or withdrawn from participation were excluded. Association analysis for the cell counts/ratios was restricted to samples with complete data on all 8 primary cell count measures. Based on the available data, 467,258 people were included in the analysis, with 18,919 migraine cases (including 4,580 incident cases) and 448,339 controls. The age sensitivity analysis (age<50) was done with 1,267 incident cases and 104,233 controls. Smoking and alcohol use were self-reported on the baseline questionnaire and categorized as current, previous, or never. The UKBB was approved by the North West Haydock Research Ethics Committee (16/NW/0274) and all patients gave informed consent. Our study is part an approved application to the UKBB (Project ID 28525).

#### Blood Processing

Blood samples were collected into 4ml EDTA tubes and stored at 4°C. The samples were transported overnight in temperature-controlled containers to the UK Biocenter (Stockport, UK) where blood cell counts were taken using Beckman Coulter LH600 Series hematology analyzers.

#### Statistical Analysis

Logistic regression was used to test associations between cell counts/ratios and prevalent migraine outcome adjusted for age and sex. Cell counts and log-transformed cell count ratios were standardized in all analyses to facilitate comparisons across measures. This was the main/simple model for the study, as similar data was available in all three cohorts and maximized the case and control sample size. Multiple secondary analyses were done, including a full model (adjusted for age (standardized), sex, body mass index (BMI, standardized), smoking, and alcohol use), ancestry-stratified, sex-stratified, sex-interaction, age<50, and incident case analyses. Incident case was defined by a migraine case with the first event date later than the baseline. The age sensitivity analysis (age<50) was done since having a first migraine in older age is rare and could reflect some missed events at earlier ages. Notably, UKBB medical records in many cases cover decades of early ages of onset. The ancestry analysis included five ancestry groups - European (EUR), African (AFR), Hispanic (AMR), South Asian (SAS), and East Asian (EAS), inferred from KING software^[17]^ with array genotype data.

### NHANES

#### Participant Selection

NHANES is a national survey of health and nutrition data from about 5,000 children and adults each year in the United States. Participants are randomly selected based on address to get a sample representative of the national population. Those who choose to participate undergo health interviews, a mobile exam center visit, and laboratory tests. We combined data from the 1991-2000, 2001-2002, and 2003-2004 exams, resulting in a total of 1,887 self-reported migraine cases and 7,242 controls. This study was approved by the NHANES Ethics Review Board, and all patients gave informed consent.

#### Blood Processing

Blood samples were drawn into 3- or 4-mL K2 EDTA tubes and processed on the Beckman Coulter DxH 800 instrument in the mobile exam center.

#### Statistical Analysis

We used simple logistic regression adjusted for age and sex to test the associations between cell counts/ratios and prevalent migraine outcome as done in UKBB. The full model was not analyzed as the smoking and alcohol data were ascertained differently than in the UKBB and FHS. Incident cases were not analyzed as the NHANES studies were cross-sectional and event timing and prospective follow-up were not available. A sex-stratified analysis was the only secondary analysis performed. An ancestry-analysis could not be done due to the large amount of missing data for self-reported race/ethnicity.

### Framingham Heart Study

#### Participant Selection

The Framingham Heart Study (FHS) is a multi-generational longitudinal cohort study. The participant data included in this study are from the Third Generation (Gen 3, N=4,095), New Offspring Spouse (NOS, N=103) and OMNI 2 (N=410) cohorts, and the Offspring (OS, N=5,124) and OMNI 1 (N=506) cohorts^[18–20]^.

Cell count and prevalent migraine status was determined through clinical diagnostic interview at OS Exam 9/OMNI 1 Exam 4 (2011-2014) and Gen 3/NOS/OMNI 2 Exam 2 (2008-2011). Incident migraine status, also from clinical diagnostic impression, was obtained from OS Exam 10/OMNI 1 Exam 5 (2019-2022) and Gen 3/NOS/OMNI 2 Exam 3 (2016-2019). Incident cases were those who reported migraine at a later Exam without migraine at the earlier Exam.

The migraine data was taken from a questionnaire (yes/no/maybe self-report) with answers judged by a physician after follow-up interview. Anyone entered as *maybe* was excluded from the analysis. There were 395 prevalent migraine cases (including 229 incident cases) and 3,172 controls for migraine-cell counts/ratios analysis. Smoking and alcohol use were self-reported and classified as current, previous, or never. This study was approved by the Boston University Institutional Review Board, and all participants gave informed consent.

#### Blood Processing

Blood was drawn from participants in a supine position after a >10 hour overnight fast into EDTA (K3) tubes (15%, Tyco Monoject). CBCs were measured using the Coulter HmX Hematology Analyzer (Beckman Coulter). A more detailed protocol has been previously described^[21]^.

#### Statistical Analysis

A generalized linear mixed effects model (implemented in lme4 R package, https://cran.r-project.org/web/packages/lme4/index.html) with logit link accounting for FHS familial correlation was used to test associations between the cell count/ratios. The simple and the full-adjustment models were considered with the additional adjustment of Exam and the time difference between Exams. Ancestry-stratified analysis was not considered due to limited non-European ancestry samples.

#### Discovery and replication analysis

In our study, UKBB was used as a discovery cohort, NHANES and FHS were used for replication for the main analysis. A False Discovery Rate (FDR) threshold of 0.05 was used for multiple testing correction in the discovery stage. A random effects model meta-analysis implemented in *metafor* R package (https://cran.r-project.org/web/packages/metafor/index.html) was used to combine FHS and NHANES results from the main model (adjusted for age and sex). Similarly, a random effects meta-analysis was performed to combine UKBB, FHS and NHANES to examine overall consistency and heterogeneity across the three cohorts. A full outline of the methods can be seen in **Figure 1**.

**Figure 1:**
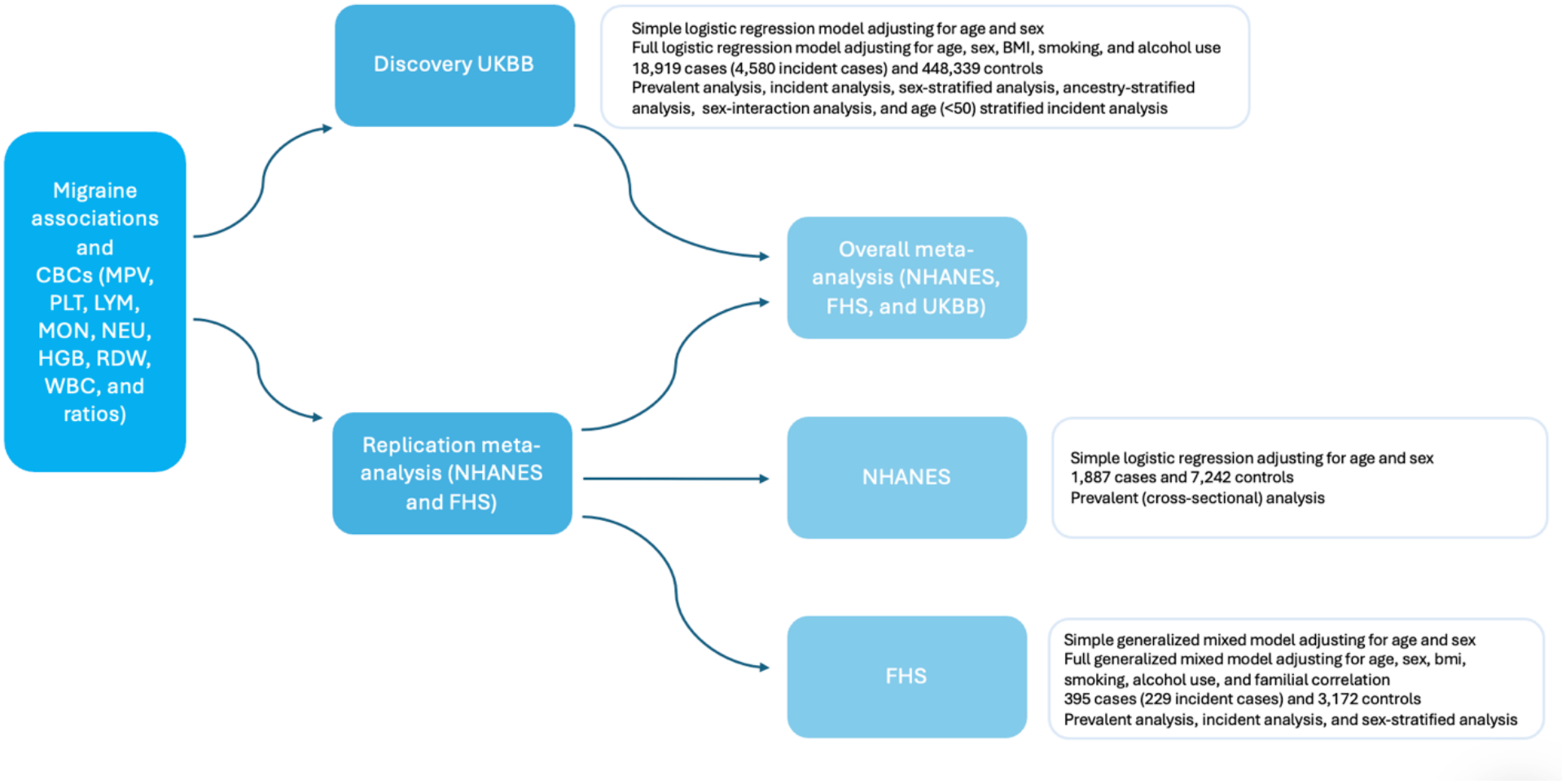
Analyses workflow

### Data Availability

UKBB data are available by application (www.ukbiobank.ac.uk). FHS data are available in dbGaP (accession number phs000007). NHANES data are publicly available (wwwn.cdc.gov/nchs/nhanes).

## Results

The demographics of the UKBB and FHS (**Table 1**) are similar. In both datasets, the average age of the cases and controls is in the mid-50s. Likewise, in both, men make up ∼50% and ∼25% of the controls and cases, respectively. BMI is similar in cases and controls. The UKBB has a higher proportion of smokers (current or previous) or alcohol drinkers (current or previous) than FHS. The UKBB ancestry demographics are similar between migraine cases and controls, the large majority being of European ancestry. Overall, FHS is also primarily of European ancestry, though the smaller Omni cohorts are primarily of non-European ancestry. NHANES (**Table 1**) is a younger cohort, especially the migraine cases, and has a slightly higher proportion of men who have migraine. NHANES represents single visit random population samples in the US and information was not available at the same level of detail as FHS and UKBB regarding past or current smoking or alcohol intake or ancestry make-up.

**Table 1:** Demographic information for all cohorts (based on prevalent analysis samples). n/a: indicates variables were unavailable or did not meet required information to include in the study.

| Cohort | UKBB |  |  | FHS |  |  | NHANES |  |  |
|---|---|---|---|---|---|---|---|---|---|
|  | Cases | Controls | p-value | Cases | Controls | p-value | Cases | Controls | p-value |
| N (incident cases) | 18919 (4580) | 448339 |  | 395 (229) | 3172 |  | 1887 | 7242 |  |
| Age<br>Q1-Q3 | 55.4 (8.0)<br>49-62 | 56.6 (8.1)<br>50-63 | 3.5E-80 | 51.7 (12.9)<br>42-62 | 55.1 (13.2)<br>45-65 | 2.2E-6 | 43.9 (16.5)<br>30-55 | 51.5 (19.3)<br>34-68 | 4.7E-55 |
| Male sex | 4468 (23.6%) | 209912 (46.8) | <4.9E-324 | 98 (24.8%) | 1592 (50.2%) | 4.0E-20 | 598 (31.7%) | 3761 (51.9%) | 1.0E-53 |
| MPV (fL)<br>Q1-Q3 | 9.33 (1.08)<br>8.59-9.93 | 9.33 (1.08)<br>8.59-9.95 | 0.73 | 8.64 (0.93)<br>8.00-9.10 | 8.65 (0.97)<br>8.00-9.20 | 0.95 | 8.28 (0.91)<br>7.60-8.80 | 8.24 (0.88)<br>7.60-8.80 | 0.09 |
| PLT (10<sup>9</sup>/L)<br>Q1-Q3 | 262.9 (61.7)<br>221.9-298.0 | 252.7 (59.9)<br>213.0-286.7 | 5.6E-117 | 252.6 (61)<br>212.0-290.0 | 241.0 (56)<br>203.0-272.0 | 1.7E-4 | 273 (70.4)<br>226.0-312.0 | 259.5 (66.9)<br>214.00-296.0 | 1.8E-14 |
| LYM (10<sup>9</sup>/L)<br>Q1-Q3 | 1.98 (1.14)<br>1.53-2.30 | 1.96 (1.12)<br>1.50-2.30 | 0.04 | 1.69 (0.52)<br>1.30-2.00 | 1.63 (0.55)<br>1.30-1.90 | 0.050 | 2.20 (0.77)<br>1.70-2.60 | 2.10 (1.52)<br>1.60-2.40 | 4.2E-3 |
| MON (10<sup>9</sup>/L)<br>Q1-Q3 | 0.46 (0.21)<br>0.34-0.53 | 0.48 (0.22)<br>0.37-0.57 | 1.5E-32 | 0.50 (0.15)<br>0.40-0.60 | 0.51 (0.15)<br>0.40-0.60 | 0.15 | 0.55 (0.19)<br>0.40-0.60 | 0.56 (0.20)<br>0.40-0.70 | 0.09 |
| NEU (10<sup>9</sup>/L)<br>Q1-Q3 | 4.18 (1.39)<br>3.22-4.90 | 4.23 (1.41)<br>3.27-4.97 | 7.6E-6 | 3.55 (1.25)<br>2.70-4.15 | 3.53 (1.19)<br>2.70-4.10 | 0.56 | 4.58 (1.91)<br>3.20-5.50 | 4.36 (1.74)<br>3.20-5.20 | 2.6E-6 |
| HGB (g/dL)<br>Q1-Q3 | 13.81 (1.18)<br>13.05-14.56 | 14.19 (1.24)<br>13.35-15.05 | <4.9E-324 | 13.6 (1.17)<br>12.80-14.30 | 14.0 (1.22)<br>13.20-14.90 | 2.0E-11 | 14.0 (1.55)<br>13.00-15.00 | 14.4 (1.51)<br>13.40-15.40 | 5.7E-23 |
| RDW (%)<br>Q1-Q3 | 13.49 (0.99)<br>12.90-13.88 | 13.49 (0.98)<br>12.90-13.86 | 0.55 | 13.0 (0.84)<br>12.40-13.30 | 12.9 (0.87)<br>12.40-13.30 | 0.68 | 12.8 (1.19)<br>12.1-13.10 | 12.8 (1.13)<br>12.10-13.10 | 0.45 |
| WBC (10<sup>9</sup>/L)<br>Q1-Q3 | 6.83 (2.03)<br>5.60-7.80 | 6.88 (2.03)<br>6.65-7.85 | 2.8E-4 | 5.94 (1.60)<br>4.80-6.70 | 5.89 (1.53)<br>4.80-6.70 | 0.35 | 7.58 (2.31)<br>5.90-8.90 | 7.27 (2.54)<br>5.70-8.40 | 1.7E-6 |
| BMI<br>Q1-Q3 | 27.26 (5.05)<br>23.74-29.79 | 27.41 (4.76)<br>24.15-29.87 | 9.0E-6 | 28.2 (5.91)<br>24.06-31.28 | 28.2 (5.44)<br>24.41-31.00 | 0.93 | n/a |  |  |
| Never smoker<br>Previous smoker<br>Current smoker | 11174 (59.1%)<br>6103 (32.2%)<br>1642 (8.7%) | 244465 (54.5%)<br>156396 (34.9%)<br>47478 (10.6%) | 6.5E-37 | 355 (89.9%)<br>13 (3.3%)<br>27 (6.8%) | 2859 (90.1%)<br>79 (2.5%)<br>234 (7.4%) | 0.67 | n/a |  |  |
| AFR<br>AMR<br>EAS<br>EUR<br>SAS<br>Mixed | 254 (1.3%)<br>50 (0.3%)<br>50 (0.3%)<br>18189 (96.1%)<br>280 (1.5%)<br>96 (0.5%) | 8008 (1.8%)<br>875 (0.2%)<br>2362 (0.5%)<br>426485 (95.1%)<br>8424 (1.9%)<br>2185 (0.5) | 1.0E-12 | 4 (1.1%)<br>10 (2.6%)<br>2 (0.5%)<br>359 (95.0%)<br>1 (0.3%)<br>2 (0.5%) | 63 (2.1%)<br>83 (2.8%)<br>39 (1.3%)<br>2766 (92.4%)<br>20 (0.7%)<br>23 (0.8%) | 0.30 | n/a |  |  |
| Never drinks<br>Previous drinker<br>Current drinker | 1321 (7.0%)<br>1254 (6.6%)<br>16344 (86.4%) | 19027 (4.2%)<br>15402 (3.4%)<br>413910 (92.3%) | 1.0E-189 | 42 (10.6%)<br>43 (10.9%)<br>310 (78.5%) | 291 (9.2%)<br>313 (9.9%)<br>2568 (81.0%) | 0.45 | n/a |  |  |

### Prevalent Migraine Case Analysis

#### UKBB

We performed a simple (age-sex adjusted) prevalent analysis on the UKBB data using all migraine cases (**Table 2**, **Figure 2**). Out of the 17 traits examined, 10 were associated with a difference in the prevalence of migraine, including PLT count (OR=1.058, 95% CI [1.043, 1.073], p=2.09E-14). Other traits that were significant included MPV.PLT (OR=0.949, 95% CI [0.935,0.964], p=1.51E-11) and MPV (OR=0.976, 95% CI [0.962, 0.991], p=0.0013). HGB, NEU and WBC were also significant.

**Figure 2:**
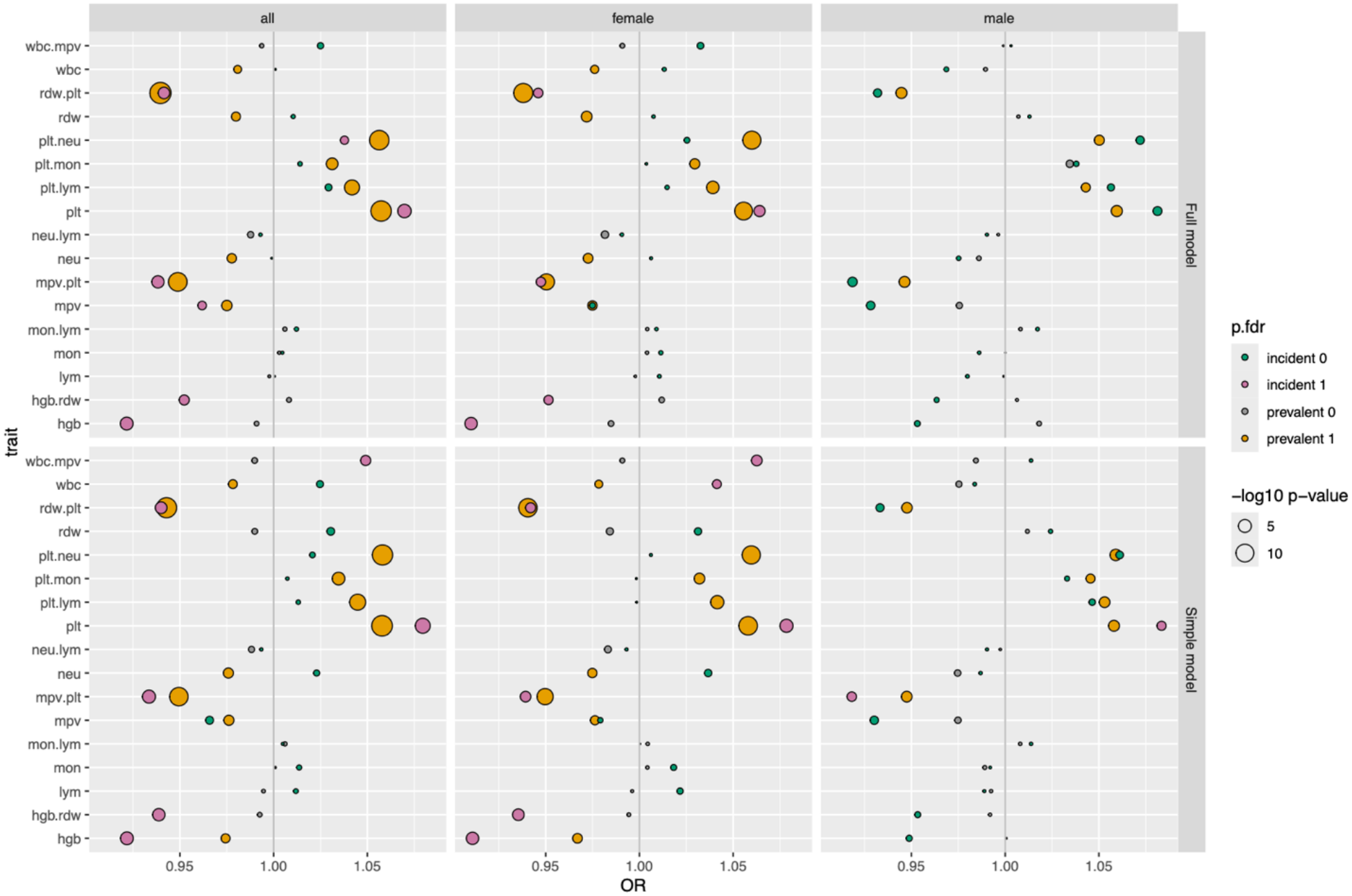
Simple (age and sex adjusted) and full (age, sex, BMI, smoking, and alcohol use adjusted) model UKBB overall and sex-stratified results

**Table 2:** UKBB simple (age and sex adjusted) and full (age, sex, BMI, smoking, and alcohol use adjusted) model overall prevalent results. Boldfaced p-values indicate signals that passed the FDR 0.05 threshold.

| **Prevalent analysis** | **UKBB (simple): N=467,258** |  |  | **UKBB (full): N=467,258** |  |  |
|---|---|---|---|---|---|---|
| **Trait** | **OR** | **OR.95ci** | **p** | **OR** | **OR.95ci** | **p** |
| **MPV.PLT** | 0.949 | (0.935,0.964) | **1.51E-11** | 0.949 | (0.935,0.963) | **8.51E-12** |
| **PLT.LYM** | 1.045 | (1.029,1.061) | **9.81E-09** | 1.042 | (1.026,1.058) | **1.19E-07** |
| **PLT.MON** | 1.035 | (1.019,1.051) | **1.46E-05** | 1.031 | (1.015,1.047) | **9.41E-05** |
| **PLT.NEU** | 1.058 | (1.043,1.074) | **6.99E-14** | 1.056 | (1.041,1.072) | **7.47E-13** |
| **HGB.RDW** | 0.993 | (0.977,1.008) | 0.35 | 1.008 | (0.992,1.024) | 0.31 |
| **RDW.PLT** | 0.943 | (0.928,0.958) | **8.91E-14** | 0.940 | (0.925,0.954) | **2.79E-15** |
| **NEU.LYM** | 0.988 | (0.974,1.003) | 0.12 | 0.988 | (0.973,1.003) | 0.10 |
| **MON.LYM** | 1.006 | (0.991,1.022) | 0.44 | 1.006 | (0.991,1.021) | 0.44 |
| **WBC.MPV** | 0.990 | (0.976,1.004) | 0.17 | 0.994 | (0.979,1.009) | 0.41 |
| **MPV** | 0.976 | (0.962,0.991) | **0.0013** | 0.975 | (0.961,0.989) | **7.55E-04** |
| **PLT** | 1.058 | (1.043,1.073) | **2.09E-14** | 1.057 | (1.042,1.073) | **3.71E-14** |
| **LYM** | 0.995 | (0.977,1.012) | 0.55 | 0.998 | (0.981,1.015) | 0.79 |
| **MON** | 1.001 | (0.986,1.016) | 0.91 | 1.003 | (0.988,1.018) | 0.69 |
| **NEU** | 0.976 | (0.961,0.990) | **0.0013** | 0.978 | (0.963,0.993) | **0.0039** |
| **HGB** | 0.974 | (0.956,0.993) | **0.0059** | 0.991 | (0.973,0.993) | 0.34 |
| **RDW** | 0.990 | (0.976,1.004) | 0.16 | 0.980 | (0.966,0.994) | **0.0056** |
| **WBC** | 0.978 | (0.963,0.994) | **0.0072** | 0.981 | (0.965,0.997) | **0.0227** |

In the simple (age adjusted) sex-stratified analyses (**Supplementary Materials; eTable 1**, **Figure 2**), the effects were similar in both males and females and were aligned with the combined analyses. The ancestry-stratified analysis (age-sex adjusted) (**Supplementary Materials; eTable 2, eFigure 1**), yielded similar results for the European (EUR) ancestry individuals to the findings in the overall cohort, including MPV.PLT (OR=0.952, 95% CI [0.937, 0.967], p=4.18E-10), MPV (OR=0.978, 95% CI [0.964, 0.993], p=0.0042), PLT (OR=1.056, 95% CI [1.040, 1.071], p=6.24E-13), and HGB (OR=0.967, 95% CI [0.948, 0.985], p=4.90E-4). Results in other ancestry groups of smaller samples were often not strictly significant -- although South Asian (SAS) ancestry has four significant signals in both the simple and full models -- but had consistent effect directions to the EUR analysis.

In secondary analyses, we also performed a full (age, sex, BMI, smoking, and alcohol use adjusted) prevalent analysis using all migraine cases (**Table 2**, **Figure 2**). Findings were similar to the simple models, suggesting limited confounding by these additional factors. The full model sex-stratified analyses (**Supplementary Materials; eTable 1**, **Figure 2**) had similar patterns to the overall analyses (simple and full models). We performed a sex-interaction analysis using the full model (**Supplementary Materials; eTable 3**), and none of the interaction terms were significant. This is consistent with the overall pattern of similar results across sexes (**Figure 2**) suggesting there is no formal sex interaction. In the full model ancestry analyses (**Supplementary Materials; eTable 4**, **Figure 2**), results in the EUR group were similar to the overall findings. SAS ancestry also had significant results in NEU, MPV, WBC and PLT.NEU.

The similarity in results between the simple and fully-adjusted models, sex-specific models, and ancestry-specific models suggests that these covariate adjustments or stratifications have little effect on the main significant associations between the core cell counts/ratios and migraine risk. This supports the use of the simple model applied in FHS and NHANES prevalent analyses, where some covariates were not available, in order to replicate the UKBB findings.

#### CBC Replication and Meta-Analysis

The NHANES-FHS meta-analysis served as a replication cohort and the results were directionally consistent with observations in UKBB, including PLT count (OR=1.074, 95% CI [1.025,1.126], p=0.0030 in overall sample (**Table 3**); OR=1.087, 95% CI [1.028,1.150], p=0.0035 in female) in the simple (age and sex adjusted) model (**Supplementary Materials; eTable 5**). These estimates are comparable to those from the joint meta-analysis and consistent with a lack of heterogeneity across the 3 cohorts (**Table 3**). Similar results were seen for FHS and NHANES in individual analyses (**eTable6**).

**Table 3:**
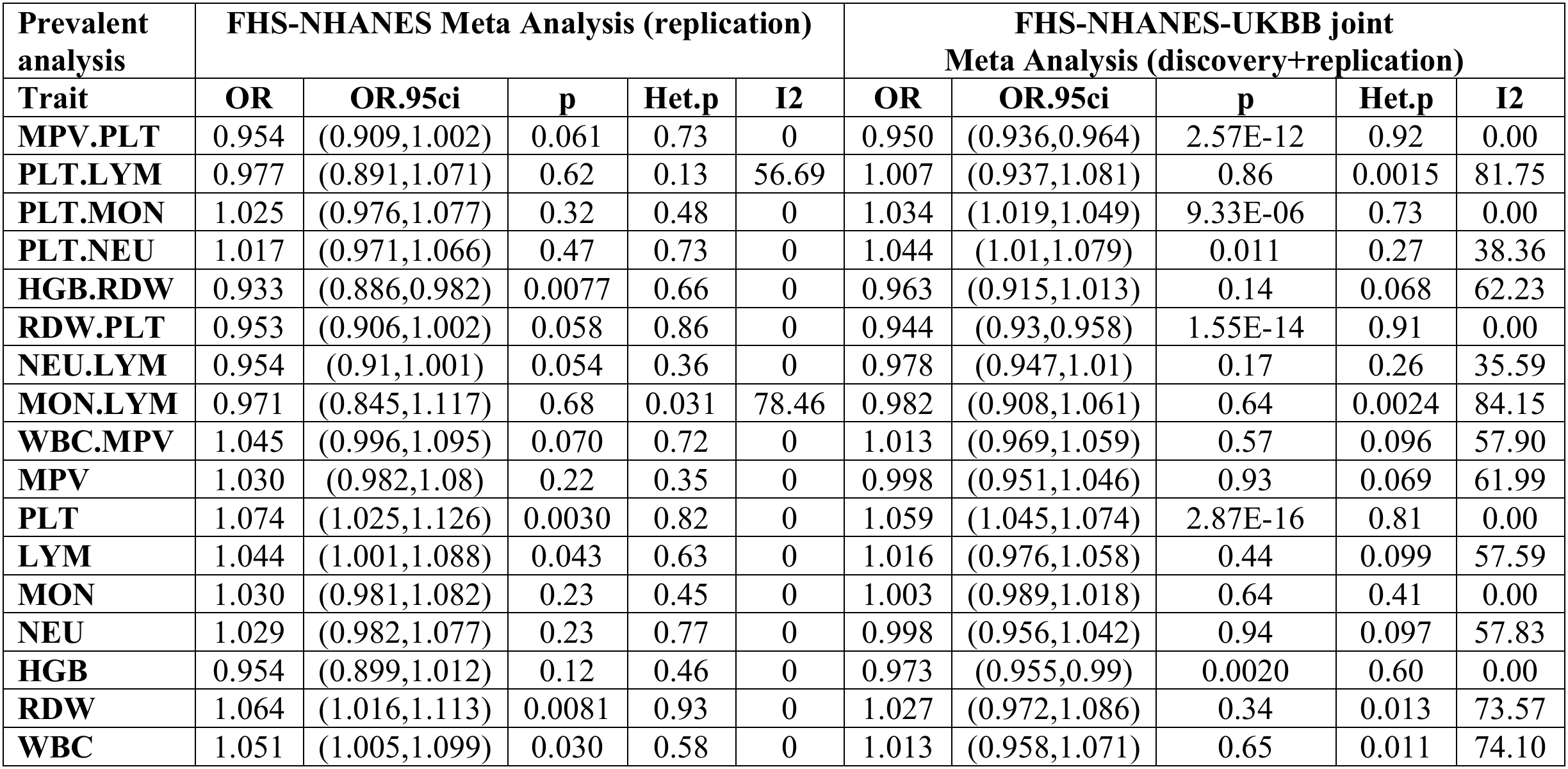
FHS-NHANES Meta Analysis simple (age and sex adjusted) model overall prevalent results, along with joint UKBB-FHS-NHANES (age and sex adjusted) meta-analysis results.

#### Incident Migraine Case Analysis

The UKBB had many significant associations in the full (age, sex, BMI, smoking, and alcohol use adjusted) models for the overall, sex-stratified, and ancestry analyses examining incident migraine (excluding baseline migraine cases). Results of significance include PLT count (OR=1.07, 95% CI [1.040, 1.101], p=3.23E-6 for overall sample; OR=1.064, 95% CI [1.030, 1.100], p=2.21E-4 for female sample), HGB (OR=0.922, 95% CI [0.889, 0.956], p=1.03E-5 for overall sample; OR=0.910, 95% CI [0.871, 0.951], p=2.56E-5 for female sample), and MPV.PLT (OR=0.938, 95% CI [0.911, 0.967], p=3.20E-5 for overall sample; OR=0.947, 95% CI [0.914, 0.982], p=0.0029 for female sample), all of which passed the multiple testing threshold. The results for the overall and sex-stratified analyses (**Supplementary Materials; eTable 7**) are consistent with each other. The overall (76.4% female) and female results are very similar, with the data having similar significance and odds ratio direction across the two analyses. None of the male results are significant, but they follow the same effect directions as the female results.

Incident case associations according to ancestry strata (full model) are more variable as expected due to smaller sample sizes and low power (**Supplementary Materials; eTable 8**) in non-European groups. The overall and European groups are very similar, with almost all results having the same effect size and direction. All significant results in the EUR analysis are significant in the overall multi-ancestry analysis, including PLT count (OR=1.069, 95% CI [1.038,1.101], p=9.08E-6), HGB (OR=0.917, 95% CI [0.883, 0.953], p=8.55E-6), and MPV.PLT (OR=0.937, 95% CI [0.909, 0.967], p=4.70E-5). Nothing is significant alone in the African, Hispanic, East Asian, or South Asian groups, most likely due to the small sample sizes and low power, but the odds ratios tend to all be in the same direction (e.g., for PLT, OR=1.122, 95% CI [0.944,1.332], p=0.19 in AFR; OR=1.301, 95% CI [0.882,1.919], p=0.19 in AMR; OR=1.047, 95% CI [0.873,1.256], p=0.62 in SAS; OR=1.362, 95% CI [0.841,2.204], p=0.21 in EAS. For HGB, AFR (OR=0.869, 95% CI [0.709,1.064], p=0.17) and AMR (OR=0.83, 95% CI [0.515,1.347], p=0.46) ancestry groups have odds ratios in the same direction as EUR and overall results. However, EAS (OR=1.078, 95% CI [0.596,1.952], p=0.80) and SAS (OR=1.112, 95% CI [0.887,1.393], p=0.36) ancestry have HGB in the opposite direction, although not significant. We also did a full model sex-interaction analysis (**Supplementary Materials; eTable 8**) and none of the results were significant, meaning there is no statistical evidence that supports sex interaction. In the full model age-sensitivity analysis (age<50), we still observed signals (FDR threshold of 0.05), though weakened in MPV.PLT, RDW.PLT, PLT, and HGB with similar effect sizes. Prospective events in NHANES were not available precluding any analysis.

Despite limited sample sizes and only several years of follow-up, we analyzed incident cases in FHS using the full model. There were no significant associations in all analyses (**Supplementary Materials; eTable 10**). However, PLT demonstrated a directionally consistent effect with the UKBB and nearly reached nominal significance among females (OR=1.165. 95% CI [0.992, 1.369], p=0.06).

## Discussion

Increased platelet count appears to be associated with an increased prevalence of migraine. It was also strongly associated with migraine in the overall incident analysis and seems to underlie the significant results for many of the ratios measured which include platelet count. Results were fairly consistent across men and women, although lower sample size among men may have limited the ability to detect statistically significant effects in this subgroup. Results were also relatively similar across ancestry groups, although power was limited in many of the subgroups. In the UKBB, we also observed evidence that increased hemoglobin, neutrophil counts or total white blood cell counts were associated with decreased migraine prevalence, but these findings were not replicated in all the UKBB secondary analyses and were not replicated in the NHANES-FHS meta-analysis.

Migraine is known to have sex differences in prevalence, with women making up most migraine cases. This difference is first seen during puberty, when the prevalence of migraine increases in both sexes, but more so in women^[22^ ^23].^ This divergence continues throughout life, but prevalence does decrease with age in both sexes^[22]^. Many women experience menstrual related migraine, and migraine frequency and severity may improve with pregnancy and menopause^[23]^. Interestingly, platelet count appears relatively similar in males and females until around the age of fourteen, and then females start to have higher platelet counts, suggesting that puberty might affect platelet production^[24]^. The onset of puberty in females is marked by an increase in estrogen. Estrogen is also thought to influence platelet count^[25^ ^26]^, specifically by increasing it, as estrogen signaling is known to promote hematopoiesis and erythropoiesis^[27]^. It is possible that the increase in platelet count that occurs in females during puberty could be connected to the increased prevalence of migraine that occurs at about the same time. In our study however, we observed higher platelet counts were associated with increased migraine prevalence in both women and men, suggesting that sex-differences in migraine and platelet counts alone do not fully account for the association between them.

Other migraine studies have also observed associations between higher platelet count and migraine. Platelet count and migraine with aura were positively, but weakly, associated in the population-based Rotterdam study^[10]^. Another study looking at platelet count during a migraine attack found that individuals with migraine had significantly higher platelet counts when compared to healthy controls^[28]^. In a comparison of platelet counts in migraine and tension-type headaches, those with migraine had higher platelet counts than those with tension-type headaches^[13]^. In contrast, other studies comparing migraine and tension-type headaches found that platelet counts were higher in those with tension-type headaches^[29^ ^30]^. However, study populations in the latter were composed of children and/or adolescents, limiting direct comparison to ours and other studies. Despite this, the findings point to a general idea of higher platelet count playing a role in headaches and headache development, which would encompass migraine.

Essential thrombocythemia (ET) is a medical condition characterized by an overproduction of platelets and an extremely high platelet count. Interestingly, one of the common manifestations of this disorder is headaches, more specifically migraine with aura-like headaches^[11^ ^31]^. Antiaggregating and antiplatelet medications have been shown to reduce the frequency and severity of the migraine-like headaches or even stop them in people with ET^[11^ ^32]^. Low-dose aspirin has also been shown to have a very modest effect on reducing migraine frequency and other related characteristics^[33]^, lending more support to the idea that antiplatelet medications can be used for migraine. These observations seem to point to platelets and high platelet count being involved in migraine pathology, which is in line with the associations seen in our data. Although ET is a very extreme case of high platelet count, the associations seen lend their support to increased platelet count playing a role in the development and onset of migraine.

A few studies did not observe an association between platelet count and migraine^[34^ ^35]^. However, differences in sample sizes, which tend to be small, participant selection (headache clinic vs emergency department vs population-based study), whether or not a healthy control population is included, types of headache included (migraine in general, migraine with and without aura, tension-type), and whether or not the participant is experiencing a migraine attack during the blood draw may explain some of the difference seen across studies. In contrast, our study was much larger and more well-powered to detect more modest associations.

This study has several limitations. A majority of the migraine data was self-reported, and we lack information on duration, frequency, or subtype of migraine. This extends into the incident cases, as we are unable to determine whether or not these cases are truly new reports from people who have never previously reported migraine. Several studies have shown differences in platelet activity based on subtype, but this could not be included in this analysis. Our sample was primarily composed of individuals of European ancestry so the generalizability to other groups may be limited. However, among the population studies to date of platelet count/MPV this study also ranks as one of the largest to our knowledge.

Our results support the hypothesis that platelets may have a contributory role in the development of migraine among both men and women. However, elucidation of more precise roles of platelets in migraine and their utility as one component of prediction of migraine attacks will require large, prospective studies with both repeated platelet measurements and more detailed recording of migraine and headache recurrence and symptomatic subtyping.

## GLOSSARY

NSAIDs: Nonsteroidal anti-inflammatory drugs
CBC: complete blood cell count
UKBB: UK Biobank
NHANES: National Health and Nutrition Examination Survey
FHS: Framingham Heart Study
BMI: body mass index
NOS: New Offspring Spouse
OS: Offspring
FDR: false discovery rate
US: United States
OR: odds ratio
CI: confidence interval
HSC: hematopoietic stem cell
Erα: estrogen receptor alpha
IDA: iron deficiency anemia
ET: essential thrombocythemia
MPV: mean platelet volume
PLT: platelet count
LYM: lymphocyte count
MON: monocyte count
NEU: neutrophil count
HGB: hemoglobin
RDW: red cell distribution width
WBC: white blood cell count
AFR: African
AMR: Hispanic
EAS: East Asian
EUR: European
SAS: South Asian

## Author Contributions

E. Sjodin: drafting / revision of the manuscript for content; study concept or design; analysis or interpretation of data. D. Chasman: drafting / revision of the manuscript for content; study concept or design analysis or interpretation of data. P. Rist: drafting / revision of the manuscript for content; study concept or design analysis or interpretation of data. M-H. Chen: drafting / revision of the manuscript for content; study concept or design analysis or interpretation of data. A.D. Johnson: drafting / revision of the manuscript for content; major role in the acquisition of data; study concept or design analysis or interpretation of data.

## Data Availability

https://wwwn.cdc.gov/nchs/nhanes/

## References

1. Danese E, Montagnana M, Lippi G. Platelets and migraine. Thromb Res 2014;134(1):17–22 doi: 10.1016/j.thromres.2014.03.055 [published Online First: 20140412].

2. Tietjen GE, Khubchandani J. Platelet dysfunction and stroke in the female migraineur. Curr Pain Headache Rep 2009;13(5):386–91 doi: 10.1007/s11916-009-0063-4.

3. Zeller JA, Frahm K, Baron R, Stingele R, Deuschl G. Platelet-leukocyte interaction and platelet activation in migraine: a link to ischemic stroke? J Neurol Neurosurg Psychiatry 2004;75(7):984–7 doi: 10.1136/jnnp.2003.019638.

4. Sarchielli P, Gallai V. Platelets in migraine. The Journal of Headache and Pain 2001;2(1):s61–s66 doi: 10.1007/s101940170012.

5. Gasparini CF, Smith RA, Griffiths LR. Genetic and biochemical changes of the serotonergic system in migraine pathobiology. J Headache Pain 2017;18(1):20 doi: 10.1186/s10194-016-0711-0 [published Online First: 20170213].

6. Yazar HO, Yazar T, Aygun A, Kaygisiz S, Kirbas D. Evaluation of simple inflammatory blood parameters in patients with migraine. Ir J Med Sci 2020;189(2):677–83 doi: 10.1007/s11845-019-02136-y [published Online First: 20191122].

7. Saricam G. Relationship between migraine headache and hematological parameters. Acta Neurol Belg 2021;121(4):899–905 doi: 10.1007/s13760-020-01362-x [published Online First: 20200428].

8. Lee SH, Kim JH, Kwon YS, Sohn JH. Role of Peripheral Inflammatory Markers in Patients with Acute Headache Attack to Differentiate between Migraine and Non-Migraine Headache. J Clin Med 2022;11(21) doi: 10.3390/jcm11216538 [published Online First: 20221103].

9. Couch JR, Hassanein RS. Platelet aggregability in migraine. Neurology 1977;27(9):843–8 doi: 10.1212/wnl.27.9.843.

10. Acarsoy C, Ruiter R, Bos D, Ikram MK. No association between blood-based markers of immune system and migraine status: a population-based cohort study. BMC Neurol 2023;23(1):445 doi: 10.1186/s12883-023-03496-w [published Online First: 20231218].

11. McIntyre KJ, Hoagland HC, Silverstein MN, Petitt RM. Essential thrombocythemia in young adults. Mayo Clin Proc 1991;66(2):149–54 doi: 10.1016/s0025-6196(12)60486-8.

12. Ozdemir HH, Donder A. Evaluation of Neutrophil-to-Lymphocyte Ratio, Platelet-to-Lymphocyte Ratio, and C-Reactive Protein in Tension-Type Headache Patients. J Neurosci Rural Pract 2021;12(3):566–70 doi: 10.1055/s-0041-1730124 [published Online First: 20210615].

13. Surbakti KP, Batubara CA, Mahendrayana E. Hematologic Differences Between Migraineurs and Tension-Type Headache Patients. Med Arch 2023;77(6):482–88 doi: 10.5455/medarh.2023.77.482-488.

14. Shi C, Li S, Liang Y, et al. A phenome-wide association study reveals novel phenotypic determinants of migraine risk in the UK Biobank. Headache 2026;66(7):1474–88 doi: 10.1111/head.70079 [published Online First: 20260610].

15. Bycroft C, Freeman C, Petkova D, et al. The UK Biobank resource with deep phenotyping and genomic data. Nature 2018;562(7726):203–09 doi: 10.1038/s41586-018-0579-z [published Online First: 20181010].

16. Nost TH, Alcala K, Urbarova I, et al. Systemic inflammation markers and cancer incidence in the UK Biobank. Eur J Epidemiol 2021;36(8):841–48 doi: 10.1007/s10654-021-00752-6 [published Online First: 20210525].

17. Manichaikul A, Mychaleckyj JC, Rich SS, Daly K, Sale M, Chen WM. Robust relationship inference in genome-wide association studies. Bioinformatics 2010;26(22):2867–73 doi: 10.1093/bioinformatics/btq559 [published Online First: 20101005].

18. Feinleib M, Kannel WB, Garrison RJ, McNamara PM, Castelli WP. The Framingham Offspring Study. Design and preliminary data. Prev Med 1975;4(4):518–25 doi: 10.1016/0091-7435(75)90037-7.

19. Splansky GL, Corey D, Yang Q, et al. The Third Generation Cohort of the National Heart, Lung, and Blood Institute’s Framingham Heart Study: design, recruitment, and initial examination. Am J Epidemiol 2007;165(11):1328–35 doi: 10.1093/aje/kwm021 [published Online First: 20070319].

20. Tsao CW, Vasan RS. Cohort Profile: The Framingham Heart Study (FHS): overview of milestones in cardiovascular epidemiology. Int J Epidemiol 2015;44(6):1800–13 doi: 10.1093/ije/dyv337.

21. Sloan A, Gona P, Johnson AD. Cardiovascular correlates of platelet count and volume in the Framingham Heart Study. Ann Epidemiol 2015;25(7):492–8 doi: 10.1016/j.annepidem.2015.01.010 [published Online First: 20150131].

22. Allais G, Chiarle G, Sinigaglia S, Airola G, Schiapparelli P, Benedetto C. Gender-related differences in migraine. Neurol Sci 2020;41(Suppl 2):429–36 doi: 10.1007/s10072-020-04643-8.

23. Peterlin BL, Gupta S, Ward TN, Macgregor A. Sex matters: evaluating sex and gender in migraine and headache research. Headache 2011;51(6):839–42 doi: 10.1111/j.1526-4610.2011.01900.x.

24. Biino G, Santimone I, Minelli C, et al. Age- and sex-related variations in platelet count in Italy: a proposal of reference ranges based on 40987 subjects’ data. PLoS One 2013;8(1):e54289 doi: 10.1371/journal.pone.0054289 [published Online First: 20130131].

25. Santimone I, Di Castelnuovo A, De Curtis A, et al. White blood cell count, sex and age are major determinants of heterogeneity of platelet indices in an adult general population: results from the MOLI-SANI project. Haematologica 2011;96(8):1180–8 doi: 10.3324/haematol.2011.043042 [published Online First: 20110505].

26. Segal JB, Moliterno AR. Platelet counts differ by sex, ethnicity, and age in the United States. Ann Epidemiol 2006;16(2):123–30 doi: 10.1016/j.annepidem.2005.06.052 [published Online First: 20051024].

27. Quatrini L, Ricci B, Ciancaglini C, Tumino N, Moretta L. Regulation of the Immune System Development by Glucocorticoids and Sex Hormones. Front Immunol 2021;12:672853 doi: 10.3389/fimmu.2021.672853 [published Online First: 20210623].

28. Karabulut KU, Egercioglu TU, Uyar M, Ucar Y. The change of neutrophils/lymphocytes ratio in migraine attacks: A case-controlled study. Ann Med Surg (Lond) 2016;10:52–6 doi: 10.1016/j.amsu.2016.07.023 [published Online First: 20160727].

29. Benedick A, Zeharia A, Markus TE. Comparison of Thrombocyte Count Between Pediatric Patients With Migraine or Tension-Type Headache: A Retrospective Cohort Study. J Child Neurol 2019;34(13):824–29 doi: 10.1177/0883073819862738 [published Online First: 20190718].

30. Aksoy E. Hematological parameters in pediatric patients with primary headache. Annals of Medical Research 2020;27(12) doi: 10.5455/annalsmedres.2020.01.049.

31. Frewin R, Dowson A. Headache in essential thrombocythaemia. Int J Clin Pract 2012;66(10):976–83 doi: 10.1111/j.1742-1241.2012.02986.x [published Online First: 20120814].

32. Bousser MG, Conard J. TIA’s, migraine and platelets. Headache 1987;27(9):522.

33. Bensenor IM, Cook NR, Lee IM, Chown MJ, Hennekens CH, Buring JE. Low-dose aspirin for migraine prophylaxis in women. Cephalalgia 2001;21(3):175–83 doi: 10.1046/j.0333-1024.2001.00194.x.

34. Albayrak HO, Gurbuz H, Basaran S, Yilmaz S. Assessment of inflammation with hematological parameters in patients with migraine and tension-type headache: a prospective study from a tertiary care center. Pain Manag 2022;12(2):141–48 doi: 10.2217/pmt-2021-0036 [published Online First: 20210825].

35. Pradalier A, Abuaf N, Launay JM, Vincent D. Platelet size and volume distribution measured by automated platelet analyzer. Cephalalgia 1992;12(5):321–2 doi: 10.1046/j.1468-2982.1992.1205321.x.

