## Supplemental Tables and Figures for "Higher platelet counts are associated with migraine risk across three cohorts among more than 21,000 cases"

| **Prevalent Analysis** | **UKBB (simple)** | | | | **UKBB (full)** | | | |
| --- | --- | --- | --- | --- | --- | --- | --- | --- |
| **Trait (** $\frac{\text{f}}{\text{m}}$**)** | **OR** | **OR.95ci** | **p** | **N** | **OR** | **OR.95ci** | **p** | **N** |
| **MPV.PLT** | 0.950 | (0.933,0.966) | **5.63E-9** | 252878 | 0.950 | (0.934,0.967) | **9.29E-9** | 252878 |
|  | 0.948 | (0.919,0.977) | **4.99E-4** | 214380 | 0.946 | (0.918,0.975) | **3.65E-4** | 214380 |
| **PLT.LYM** | 1.042 | (1.024,1.060) | **4.94E-6** | 252878 | 1.039 | (1.021,1.058) | **1.98E-5** | 252878 |
|  | 1.053 | (1.023,1.084) | **5.22E-4** | 214380 | 1.043 | (1.012,1.074) | **0.0054** | 214380 |
| **PLT.MON** | 1.032 | (1.014,1.050) | **4.56E-4** | 252878 | 1.030 | (1.012,1.048) | **0.0012** | 252878 |
|  | 1.046 | (1.013,1.079) | **0.0057** | 214380 | 1.034 | (1.002,1.068) | 0.0377 | 214380 |
| **PLT.NEU** | 1.060 | (1.042,1.078) | **3.10E-11** | 252878 | 1.060 | (1.042,1.079) | **4.30E-11** | 252878 |
|  | 1.059 | (1.028,1.091) | **1.49E-4** | 214380 | 1.050 | (1.019,1.082) | **0.0015** | 214380 |
| **HGB.RDW** | 0.994 | (0.977,1.012) | 0.52 | 252878 | 1.012 | (0.994,1.030) | 0.19 | 252878 |
|  | 0.992 | (0.957,1.028) | 0.66 | 214380 | 1.006 | (0.971,1.043) | 0.73 | 214380 |
| **RDW.PLT** | 0.941 | (0.924,0.957) | **1.86E-11** | 252878 | 0.938 | (0.921,0.955) | **2.45E-12** | 252878 |
|  | 0.948 | (0.919,0.977) | **6.21E-4** | 214380 | 0.945 | (0.916,0.974) | **2.91E-4** | 214380 |
| **NEU.LYM** | 0.983 | (0.966,1.001) | 0.06 | 252878 | 0.982 | (0.965,0.999) | 0.04 | 252878 |
|  | 0.997 | (0.969,1.027) | 0.86 | 214380 | 0.996 | (0.968,1.026) | 0.80 | 214380 |
| **MON.LYM** | 1.004 | (0.987,1.022) | 0.62 | 252878 | 1.004 | (0.987,1.022) | 0.64 | 252878 |
|  | 1.008 | (0.977,1.040) | 0.61 | 214380 | 1.008 | (0.977,1.040) | 0.61 | 214380 |
| **WBC.MPV** | 0.991 | (0.974,1.008) | 0.29 | 252878 | 0.991 | (0.974,1.008) | 0.30 | 252878 |
|  | 0.984 | (0.956,1.014) | 0.30 | 214380 | 0.999 | (0.969,1.030) | 0.95 | 214380 |
| **MPV** | 0.976 | (0.960,0.993) | **0.0052** | 252878 | 0.975 | (0.959,0.991) | **0.0031** | 252878 |
|  | 0.975 | (0.946,1.005) | 0.10 | 214380 | 0.976 | (0.947,1.005) | 0.11 | 214380 |
| **PLT** | 1.058 | (1.041,1.075) | **1.17E-11** | 252878 | 1.056 | (1.038,1.073) | **8.85E-11** | 252878 |
|  | 1.058 | (1.026,1.091) | **3.44E-4** | 214380 | 1.060 | (1.027,1.093) | **2.37E-4** | 214380 |
| **LYM** | 0.996 | (0.975,1.018) | 0.73 | 252878 | 0.998 | (0.976,1.020) | 0.85 | 252878 |
|  | 0.992 | (0.962,1.024) | 0.64 | 214380 | 0.999 | (0.973,1.026) | 0.95 | 214380 |
| **MON** | 1.004 | (0.988,1.021) | 0.62 | 252878 | 1.004 | (0.987,1.021) | 0.64 | 252878 |
|  | 0.989 | (0.959,1.020) | 0.49 | 214380 | 1.000 | (0.971,1.030) | 0.99 | 214380 |
| **NEU** | 0.975 | (0.958,0.992) | **0.0038** | 252878 | 0.973 | (0.956,0.990) | **0.0022** | 252878 |
|  | 0.975 | (0.946,1.004) | 0.09 | 214380 | 0.986 | (0.956,1.017) | 0.37 | 214380 |
| **HGB** | 0.967 | (0.946,0.988) | **0.0024** | 252878 | 0.985 | (0.964,1.007) | 0.17 | 252878 |
|  | 1.001 | (0.965,1.038) | 0.97 | 214380 | 1.018 | (0.982,1.056) | 0.33 | 214380 |
| **RDW** | 0.984 | (0.969,1.000) | 0.0482 | 252878 | 0.972 | (0.956,0.988) | **4.77E-4** | 252878 |
|  | 1.012 | (0.979,1.046) | 0.48 | 214380 | 1.007 | (0.974,1.041) | 0.68 | 214380 |
| **WBC** | 0.978 | (0.960,0.997) | **0.0216** | 252878 | 0.976 | (0.957,0.995) | **0.0159** | 252878 |
|  | 0.975 | (0.945,1.007) | 0.12 | 214380 | 0.990 | (0.959,1.021) | 0.51 | 214380 |

eTable 1: Prevalent sex-stratified results for UKBB simple (age and sex adjusted) and full (age, sex, BMI, smoking, and alcohol use adjusted) models. Female results are on the top half and male results are on the bottom half for each trait. Boldfaced p-values indicate signals that passed FDR 0.05 threshold.

| **Trait** | **EUR N=444674** | | | **AFR N=8262** | | | **AMR N=925** | | | **SAS N=8704** | | | **EAS N=2412** | | |
| --- | --- | --- | --- | --- | --- | --- | --- | --- | --- | --- | --- | --- | --- | --- | --- |
|  | **OR** | **OR.95ci** | **p** | **OR** | **OR.95ci** | **p** | **OR** | **OR.95ci** | **p** | **OR** | **OR.95ci** | **p** | **OR** | **OR.95ci** | **p** |
| **MPV.PLT** | 0.952 | (0.937,0.967) | **4.18E-10** | 0.949 | (0.845,1.067) | 0.39 | 0.929 | (0.693,1.245) | 0.62 | 0.886 | (0.786,0.998) | 0.05 | 0.799 | (0.588,1.085) | 0.15 |
| **PLT.LYM** | 1.040 | (1.024,1.056) | **5.30E-7** | 1.039 | (0.922,1.170) | 0.53 | 0.973 | (0.727,1.302) | 0.85 | 1.119 | (0.993,1.262) | 0.07 | 1.217 | (0.910,1.626) | 0.18 |
| **PLT.MON** | 1.035 | (1.018,1.051) | **2.44E-5** | 1.121 | (1.002,1.253) | 0.05 | 1.299 | (1.014,1.663) | 0.04 | 1.120 | (0.997,1.258) | 0.06 | 1.198 | (0.918,1.564) | 0.18 |
| **PLT.NEU** | 1.060 | (1.044,1.076) | **4.12E-14** | 1.058 | (0.948,1.179) | 0.31 | 1.186 | (0.911,1.543) | 0.21 | 1.305 | (1.153,1.477) | **2.51E-5** | 1.417 | (1.064,1.888) | 0.02 |
| **HGB.RDW** | 0.983 | (0.967,0.999) | 0.04 | 1.004 | (0.909,1.109) | 0.94 | 1.019 | (0.781,1.330) | 0.89 | 0.993 | (0.904,1.089) | 0.87 | 0.946 | (0.748,1.195) | 0.64 |
| **RDW.PLT** | 0.947 | (0.932,0.963) | **2.31E-11** | 0.907 | (0.804,1.024) | 0.12 | 0.847 | (0.619,1.159) | 0.30 | 0.930 | (0.823,1.050) | 0.24 | 0.787 | (0.576,1.075) | 0.13 |
| **NEU.LYM** | 0.982 | (0.967,0.998) | **0.0233** | 0.977 | (0.871,1.096) | 0.69 | 0.830 | (0.628,1.098) | 0.19 | 0.878 | (0.773,0.998) | 0.05 | 0.868 | (0.647,1.164) | 0.34 |
| **MON.LYM** | 1.002 | (0.986,1.018) | 0.80 | 0.916 | (0.818,1.026) | 0.13 | 0.741 | (0.572,0.959) | 0.02 | 0.978 | (0.866,1.106) | 0.73 | 0.964 | (0.722,1.286) | 0.80 |
| **WBC.MPV** | 0.987 | (0.972,1.002) | 0.09 | 1.041 | (0.930,1.166) | 0.48 | 0.845 | (0.625,1.144) | 0.28 | 0.875 | (0.772,0.991) | 0.04 | 0.890 | (0.671,1.180) | 0.42 |
| **MPV** | 0.978 | (0.964,0.993) | **0.0042** | 1.042 | (0.929,1.169) | 0.48 | 1.025 | (0.784,1.340) | 0.86 | 0.839 | (0.743,0.947) | **0.0045** | 0.905 | (0.671,1.222) | 0.52 |
| **PLT** | 1.056 | (1.040,1.071) | **6.24E-13** | 1.072 | (0.955,1.205) | 0.24 | 1.123 | (0.849,1.486) | 0.42 | 1.093 | (0.980,1.219) | 0.11 | 1.233 | (0.928,1.639) | 0.15 |
| **LYM** | 0.998 | (0.981,1.015) | 0.81 | 1.034 | (0.844,1.266) | 0.75 | 1.201 | (0.717,2.011) | 0.49 | 0.848 | (0.698,1.030) | 0.10 | 0.913 | (0.531,1.570) | 0.74 |
| **MON** | 0.999 | (0.983,1.015) | 0.89 | 1.058 | (0.937,1.194) | 0.36 | 0.647 | (0.403,1.038) | 0.07 | 0.880 | (0.746,1.038) | 0.13 | 0.912 | (0.574,1.448) | 0.69 |
| **NEU** | 0.973 | (0.958,0.988) | **3.70E-4** | 1.044 | (0.919,1.187) | 0.51 | 0.853 | (0.620,1.175) | 0.33 | 0.774 | (0.674,0.888) | **2.74E-4** | 0.849 | (0.605,1.190) | 0.34 |
| **HGB** | 0.967 | (0.948,0.985) | **4.90E-4** | 0.981 | (0.853,1.128) | 0.79 | 0.903 | (0.645,1.263) | 0.55 | 0.996 | (0.872,1.138) | 0.96 | 0.873 | (0.631,1.206) | 0.41 |
| **RDW** | 0.997 | (0.982,1.012) | 0.69 | 0.983 | (0.906,1.065) | 0.67 | 0.900 | (0.689,1.175) | 0.44 | 1.008 | (0.931,1.092) | 0.84 | 1.003 | (0.816,1.233) | 0.98 |
| **WBC** | 0.977 | (0.961,0.993) | **0.0056** | 1.068 | (0.927,1.231) | 0.36 | 0.839 | (0.580,1.215) | 0.35 | 0.761 | (0.656,0.882) | **2.78E-4** | 0.861 | (0.591,1.252) | 0.43 |

eTable 2: UKBB simple (age and sex adjusted) model prevalent ancestry-stratified results. Boldfaced p-values indicate signals that passed FDR 0.05 threshold.

| **Trait** | **OR** | **OR.95ci** | **p** | **N** |
| --- | --- | --- | --- | --- |
| **MPV.PLT** | 0.998 | (0.964,1.033) | 0.90 | 467258 |
| **PLT.LYM** | 1.011 | (0.978,1.046) | 0.52 | 467258 |
| **PLT.MON** | 1.005 | (0.970,1.042) | 0.78 | 467258 |
| **PLT.NEU** | 0.992 | (0.959,1.027) | 0.66 | 467258 |
| **HGB.RDW** | 0.991 | (0.953,1.031) | 0.66 | 467258 |
| **RDW.PLT** | 1.011 | (0.976,1.048) | 0.54 | 467258 |
| **NEU.LYM** | 1.020 | (0.986,1.055) | 0.26 | 467258 |
| **MON.LYM** | 1.010 | (0.975,1.047) | 0.57 | 467258 |
| **WBC.MPV** | 1.000 | (0.967,1.035) | 0.99 | 467258 |
| **MPV** | 0.997 | (0.964,1.033) | 0.88 | 467258 |
| **PLT** | 1.000 | (0.966,1.035) | 0.99 | 467258 |
| **LYM** | 0.995 | (0.960,1.032) | 0.80 | 467258 |
| **MON** | 0.993 | (0.960,1.027) | 0.68 | 467258 |
| **NEU** | 1.007 | (0.973,1.042) | 0.68 | 467258 |
| **HGB** | 1.027 | (0.985,1.071) | 0.21 | 467258 |
| **RDW** | 1.034 | (0.997,1.072) | 0.07 | 467258 |
| **WBC** | 1.004 | (0.968,1.040) | 0.84 | 467258 |

eTable 3: Full (age, sex, BMI, smoking, and alcohol use adjusted) UKBB prevalent sex interaction analysis

| **Trait** | **EUR N=444674** | | | **AFR N=8262** | | | **AMR N=925** | | | **SAS N=8704** | | | **EAS N=2412** | | |
| --- | --- | --- | --- | --- | --- | --- | --- | --- | --- | --- | --- | --- | --- | --- | --- |
|  | **OR** | **OR.95ci** | **p** | **OR** | **OR.95ci** | **p** | **OR** | **OR.95ci** | **p** | **OR** | **OR.95ci** | **p** | **OR** | **OR.95ci** | **p** |
| **MPV.PLT** | 0.952 | (0.937,0.967) | **4.16E-10** | 0.947 | (0.841,1.065) | 0.36 | 0.905 | (0.675,1.212) | 0.50 | 0.890 | (0.790,1.002) | 0.05 | 0.809 | (0.593,1.105) | 0.18 |
| **PLT.LYM** | 1.034 | (1.018,1.050) | **3.03E-5** | 1.035 | (0.918,1.167) | 0.57 | 0.969 | (0.721,1.302) | 0.83 | 1.121 | (0.994,1.265) | 0.06 | 1.229 | (0.917,1.645) | 0.17 |
| **PLT.MON** | 1.032 | (1.016,1.049) | **9.20E-5** | 1.119 | (1.000,1.251) | 0.05 | 1.296 | (1.010,1.662) | 0.04 | 1.119 | (0.996,1.257) | 0.06 | 1.204 | (0.925,1.567) | 0.17 |
| **PLT.NEU** | 1.061 | (1.045,1.077) | **6.56E-14** | 1.057 | (0.947,1.180) | 0.32 | 1.162 | (0.892,1.513) | 0.27 | 1.315 | (1.161,1.489) | **1.65E-5** | 1.443 | (1.082,1.925) | 0.01 |
| **HGB.RDW** | 0.996 | (0.980,1.013) | 0.67 | 1.003 | (0.908,1.108) | 0.95 | 1.050 | (0.801,1.376) | 0.72 | 0.999 | (0.909,1.097) | 0.98 | 0.948 | (0.751,1.198) | 0.66 |
| **RDW.PLT** | 0.945 | (0.930,0.960) | **2.27E-12** | 0.907 | (0.803,1.024) | 0.11 | 0.824 | (0.601,1.130) | 0.23 | 0.933 | (0.826,1.053) | 0.26 | 0.804 | (0.585,1.104) | 0.18 |
| **NEU.LYM** | 0.977 | (0.963,0.993) | **0.0037** | 0.975 | (0.869,1.093) | 0.66 | 0.841 | (0.635,1.115) | 0.23 | 0.874 | (0.769,0.993) | 0.04 | 0.861 | (0.641,1.156) | 0.32 |
| **MON.LYM** | 0.998 | (0.983,1.014) | 0.81 | 0.915 | (0.817,1.025) | 0.12 | 0.736 | (0.566,0.956) | 0.02 | 0.980 | (0.866,1.108) | 0.74 | 0.961 | (0.724,1.277) | 0.78 |
| **WBC.MPV** | 0.990 | (0.974,1.005) | 0.19 | 1.049 | (0.934,1.179) | 0.42 | 0.888 | (0.653,1.207) | 0.45 | 0.860 | (0.756,0.977) | 0.02 | 0.851 | (0.637,1.137) | 0.27 |
| **MPV** | 0.978 | (0.963,0.993) | **0.0039** | 1.041 | (0.928,1.168) | 0.49 | 0.998 | (0.761,1.309) | 0.99 | 0.837 | (0.741,0.945) | **0.0041** | 0.892 | (0.659,1.207) | 0.46 |
| **PLT** | 1.055 | (1.039,1.071) | **1.47E-12** | 1.077 | (0.957,1.211) | 0.22 | 1.146 | (0.866,1.515) | 0.34 | 1.087 | (0.973,1.213) | 0.14 | 1.206 | (0.900,1.615) | 0.21 |
| **LYM** | 1.003 | (0.987,1.019) | 0.73 | 1.048 | (0.853,1.287) | 0.66 | 1.261 | (0.748,2.126) | 0.38 | 0.835 | (0.684,1.019) | 0.08 | 0.853 | (0.488,1.491) | 0.58 |
| **MON** | 1.000 | (0.985,1.016) | 0.99 | 1.059 | (0.939,1.193) | 0.35 | 0.653 | (0.405,1.054) | 0.08 | 0.875 | (0.741,1.033) | 0.12 | 0.859 | (0.537,1.375) | 0.53 |
| **NEU** | 0.972 | (0.957,0.988) | **4.40E-4** | 1.047 | (0.919,1.193) | 0.49 | 0.886 | (0.642,1.224) | 0.46 | 0.755 | (0.655,0.870) | **9.95E-5** | 0.804 | (0.568,1.138) | 0.22 |
| **HGB** | 0.980 | (0.961,0.999) | 0.04 | 0.987 | (0.858,1.135) | 0.85 | 0.948 | (0.677,1.328) | 0.76 | 1.002 | (0.876,1.146) | 0.98 | 0.868 | (0.628,1.198) | 0.39 |
| **RDW** | 0.988 | (0.973,1.003) | 0.12 | 0.986 | (0.910,1.069) | 0.74 | 0.887 | (0.675,1.165) | 0.39 | 1.003 | (0.925,1.087) | 0.94 | 0.996 | (0.810,1.226) | 0.97 |
| **WBC** | 0.979 | (0.962,0.996) | **0.0152** | 1.078 | (0.931,1.248) | 0.31 | 0.878 | (0.604,1.277) | 0.50 | 0.735 | (0.631,0.857) | **8.17E-5** | 0.795 | (0.538,1.176) | 0.25 |

eTable 4: UKBB full (age, sex, BMI, smoking, and alcohol use adjusted) model prevalent analysis ancestry-stratified results. Boldfaced p-values indicate signals that passed FDR 0.05 threshold.

| **Prevalent Analysis** | **NHANES** | | | | **FHS** | | | | **FHS-NHANES Meta Analysis** | | | | |
| --- | --- | --- | --- | --- | --- | --- | --- | --- | --- | --- | --- | --- | --- |
| **Trait (** $\frac{\text{f}}{\text{m}}$**)** | **OR** | **OR.95ci** | **p** | **N** | **OR** | **OR.95ci** | **p** | **N** | **OR** | **OR.95ci** | **p** | **Het.p** | **I2** |
| **MPV.PLT** | 0.950 | (0.888,1.015) | 0.13 | 4770 | 0.918 | (0.797,1.057) | 0.24 | 1877 | 0.944 | (0.888,1.003) | 0.061 | 0.67 | 0 |
|  | 0.967 | (0.884,1.059) | 0.47 | 4359 | 0.962 | (0.778,1.188) | 0.72 | 1690 | 0.967 | (0.890,1.050) | 0.42 | 0.96 | 0 |
| **PLT.LYM** | 0.927 | (0.864,0.993) | 0.03 | 4770 | 1.065 | (0.926,1.225) | 0.38 | 1877 | 0.980 | (0.857,1.120) | 0.76 | 0.08 | 66.97 |
|  | 0.969 | (0.890,1.054) | 0.46 | 4359 | 0.987 | (0.805,1.210) | 0.90 | 1690 | 0.971 | (0.898,1.050) | 0.47 | 0.87 | 0 |
| **PLT.MON** | 1.028 | (0.961,1.100) | 0.42 | 4770 | 1.019 | (0.879,1.181) | 0.80 | 1877 | 1.026 | (0.966,1.091) | 0.40 | 0.91 | 0 |
|  | 1.056 | (0.963,1.159) | 0.25 | 4359 | 0.941 | (0.751,1.181) | 0.60 | 1690 | 1.039 | (0.954,1.132) | 0.38 | 0.36 | 0 |
| **PLT.NEU** | 1.026 | (0.964,1.092) | 0.42 | 4770 | 1.091 | (0.954,1.246) | 0.20 | 1877 | 1.037 | (0.980,1.097) | 0.20 | 0.42 | 0 |
|  | 1.019 | (0.930,1.117) | 0.69 | 4359 | 0.941 | (0.754,1.173) | 0.59 | 1690 | 1.007 | (0.926,1.096) | 0.87 | 0.51 | 0 |
| **HGB.RDW** | 0.975 | (0.913,1.042) | 0.46 | 4770 | 0.878 | (0.758,1.016) | 0.08 | 1877 | 0.945 | (0.859,1.039) | 0.24 | 0.20 | 39.61 |
|  | 0.878 | (0.784,0.983) | 0.02 | 4359 | 1.050 | (0.813,1.357) | 0.71 | 1690 | 0.924 | (0.789,1.083) | 0.33 | 0.21 | 36.39 |
| **RDW.PLT** | 0.928 | (0.865,0.994) | 0.03 | 4770 | 0.928 | (0.803,1.072) | 0.31 | 1877 | 0.928 | (0.871,0.987) | 0.018 | 1.00 | 0 |
|  | 0.990 | (0.904,1.084) | 0.83 | 4359 | 0.957 | (0.775,1.182) | 0.69 | 1690 | 0.985 | (0.906,1.071) | 0.72 | 0.78 | 0 |
| **NEU.LYM** | 0.926 | (0.868,0.988) | 0.02 | 4770 | 0.972 | (0.853,1.109) | 0.68 | 1877 | 0.935 | (0.882,0.990) | 0.022 | 0.51 | 0 |
|  | 0.955 | (0.872,1.046) | 0.32 | 4359 | 1.041 | (0.836,1.297) | 0.72 | 1690 | 0.967 | (0.889,1.052) | 0.44 | 0.48 | 0 |
| **MON.LYM** | 0.907 | (0.848,0.970) | 0.0047 | 4770 | 1.043 | (0.905,1.203) | 0.56 | 1877 | 0.959 | (0.838,1.097) | 0.54 | 0.082 | 67.018 |
|  | 0.913 | (0.833,1.001) | 0.05 | 4359 | 1.043 | (0.834,1.304) | 0.71 | 1690 | 0.937 | (0.846,1.038) | 0.21 | 0.28 | 13.41 |
| **WBC.MPV** | 1.050 | (0.983,1.120) | 0.15 | 4770 | 0.994 | (0.872,1.134) | 0.93 | 1877 | 1.038 | (0.979,1.101) | 0.21 | 0.47 | 0 |
|  | 1.028 | (0.941,1.123) | 0.55 | 4359 | 1.095 | (0.879,1.364) | 0.42 | 1690 | 1.037 | (0.955,1.125) | 0.39 | 0.60 | 0 |
| **MPV** | 1.048 | (0.982,1.119) | 0.16 | 4770 | 0.975 | (0.847,1.123) | 0.72 | 1877 | 1.035 | (0.976,1.098) | 0.25 | 0.36 | 0 |
|  | 1.029 | (0.942,1.123) | 0.53 | 4359 | 0.968 | (0.783,1.197) | 0.76 | 1690 | 1.020 | (0.940,1.106) | 0.64 | 0.60 | 0 |
| **PLT** | 1.083 | (1.018,1.152) | 0.01 | 4770 | 1.108 | (0.970,1.265) | 0.13 | 1877 | 1.087 | (1.028,1.150) | 0.0035 | 0.76 | 0 |
|  | 1.051 | (0.953,1.160) | 0.32 | 4359 | 1.043 | (0.839,1.297) | 0.70 | 1690 | 1.050 | (0.96,1.148) | 0.28 | 0.95 | 0 |
| **LYM** | 1.156 | (1.047,1.276) | 0.0041 | 4770 | 1.004 | (0.893,1.130) | 0.94 | 1877 | 1.081 | (0.942,1.241) | 0.26 | 0.073 | 68.82 |
|  | 1.015 | (0.955,1.079) | 0.63 | 4359 | 1.087 | (0.868,1.360) | 0.47 | 1690 | 1.020 | (0.962,1.082) | 0.51 | 0.57 | 0 |
| **MON** | 1.040 | (0.970,1.115) | 0.27 | 4770 | 1.053 | (0.912,1.216) | 0.48 | 1877 | 1.042 | (0.979,1.110) | 0.19 | 0.88 | 0 |
|  | 0.988 | (0.907,1.077) | 0.79 | 4359 | 1.081 | (0.884,1.323) | 0.45 | 1690 | 1.002 | (0.926,1.084) | 0.96 | 0.42 | 0 |
| **NEU** | 1.021 | (0.961,1.085) | 0.50 | 4770 | 0.979 | (0.860,1.115) | 0.75 | 1877 | 1.013 | (0.959,1.071) | 0.64 | 0.57 | 0 |
|  | 1.026 | (0.935,1.127) | 0.58 | 4359 | 1.079 | (0.870,1.339) | 0.49 | 1690 | 1.035 | (0.950,1.127) | 0.44 | 0.68 | 0 |
| **HGB** | 0.982 | (0.905,1.067) | 0.67 | 4770 | 0.875 | (0.731,1.047) | 0.14 | 1877 | 0.954 | (0.864,1.053) | 0.35 | 0.25 | 24.61 |
|  | 0.983 | (0.878,1.101) | 0.77 | 4359 | 1.021 | (0.791,1.317) | 0.88 | 1690 | 0.989 | (0.892,1.097) | 0.84 | 0.79 | 0 |
| **RDW** | 1.020 | (0.962,1.082) | 0.50 | 4770 | 1.090 | (0.958,1.240) | 0.19 | 1877 | 1.032 | (0.978,1.089) | 0.25 | 0.36 | 0 |
|  | 1.182 | (1.074,1.302) | 6.58E-4 | 4359 | 0.948 | (0.749,1.199) | 0.66 | 1690 | 1.088 | (0.882,1.341) | 0.43 | 0.089 | 65.48 |
| **WBC** | 1.069 | (0.997,1.146) | 0.06 | 4770 | 0.984 | (0.865,1.120) | 0.81 | 1877 | 1.045 | (0.972,1.124) | 0.23 | 0.27 | 18.15 |
|  | 1.029 | (0.955,1.109) | 0.45 | 4359 | 1.103 | (0.888,1.371) | 0.38 | 1690 | 1.037 | (0.966,1.113) | 0.32 | 0.55 | 0 |

eTable 5: Prevalent analysis simple (age and sex adjusted) model sex-stratified results for NHANES, FHS, and FHS-NHANES meta-analysis. Female values are on the top half and male values are on the bottom half for each trait.

| **Prevalent analysis** | **NHANES** | | | | **FHS** | | | |
| --- | --- | --- | --- | --- | --- | --- | --- | --- |
| **Trait** | **OR** | **OR.95ci** | **p** | **N** | **OR** | **OR.95ci** | **p** | **N** |
| **MPV.PLT** | 0.958 | (0.908,1.011) | 0.12 | 9129 | 0.937 | (0.835,1.052) | 0.27 | 3567 |
| **PLT.LYM** | 0.943 | (0.894,0.995) | 0.03 | 9129 | 1.040 | (0.928,1.166) | 0.50 | 3567 |
| **PLT.MON** | 1.033 | (0.979,1.091) | 0.24 | 9129 | 0.985 | (0.872,1.112) | 0.81 | 3567 |
| **PLT.NEU** | 1.013 | (0.963,1.066) | 0.61 | 9129 | 1.036 | (0.926,1.159) | 0.54 | 3567 |
| **HGB.RDW** | 0.938 | (0.886,0.992) | 0.03 | 9129 | 0.910 | (0.805,1.028) | 0.12 | 3567 |
| **RDW.PLT** | 0.955 | (0.904,1.009) | 0.10 | 9129 | 0.944 | (0.840,1.062) | 0.34 | 3567 |
| **NEU.LYM** | 0.945 | (0.896,0.995) | 0.03 | 9129 | 1.000 | (0.895,1.118) | 0.99 | 3567 |
| **MON.LYM** | 0.913 | (0.865,0.964) | 0.0010 | 9129 | 1.054 | (0.936,1.187) | 0.38 | 3567 |
| **WBC.MPV** | 1.049 | (0.996,1.105) | 0.07 | 9129 | 1.025 | (0.918,1.146) | 0.66 | 3567 |
| **MPV** | 1.041 | (0.988,1.097) | 0.13 | 9129 | 0.979 | (0.872,1.100) | 0.72 | 3567 |
| **PLT** | 1.072 | (1.017,1.129) | 0.0095 | 9129 | 1.087 | (0.972,1.215) | 0.14 | 3567 |
| **LYM** | 1.049 | (1.002,1.098) | 0.04 | 9129 | 1.020 | (0.922,1.129) | 0.70 | 3567 |
| **MON** | 1.022 | (0.968,1.078) | 0.43 | 9129 | 1.073 | (0.955,1.205) | 0.24 | 3567 |
| **NEU** | 1.032 | (0.981,1.085) | 0.22 | 9129 | 1.014 | (0.909,1.130) | 0.81 | 3567 |
| **HGB** | 0.964 | (0.903,1.029) | 0.27 | 9129 | 0.908 | (0.787,1.047) | 0.18 | 3567 |
| **RDW** | 1.065 | (1.012,1.120) | 0.01 | 9129 | 1.059 | (0.950,1.180) | 0.30 | 3567 |
| **WBC** | 1.057 | (1.006,1.110) | 0.03 | 9129 | 1.022 | (0.917,1.140) | 0.69 | 3567 |

eTable 6: Simple (age and sex adjusted) prevalent analysis overall results for FHS and NHANES

| **Trait** | **Overall N=452919** | | | **Male N=211194** | | | **Female N=241725** | | |
| --- | --- | --- | --- | --- | --- | --- | --- | --- | --- |
|  | **OR** | **OR.95ci** | **p** | **OR** | **OR.95ci** | **p** | **OR** | **OR.95ci** | **p** |
| **MPV.PLT** | 0.938 | (0.911,0.967) | **3.20E-5** | 0.919 | (0.868,0.972) | 0.0032 | 0.947 | (0.914,0.982) | **0.0029** |
| **PLT.LYM** | 1.029 | (0.999,1.061) | 0.06 | 1.056 | (1.000,1.116) | 0.05 | 1.015 | (0.979,1.052) | 0.42 |
| **PLT.MON** | 1.014 | (0.983,1.046) | 0.37 | 1.038 | (0.978,1.101) | 0.22 | 1.004 | (0.968,1.041) | 0.84 |
| **PLT.NEU** | 1.038 | (1.007,1.069) | **0.0151** | 1.072 | (1.014,1.133) | 0.01 | 1.025 | (0.989,1.063) | 0.17 |
| **HGB.RDW** | 0.952 | (0.924,0.981) | **0.0013** | 0.963 | (0.904,1.027) | 0.26 | 0.952 | (0.920,0.984) | **0.0040** |
| **RDW.PLT** | 0.942 | (0.913,0.971) | **1.20E-4** | 0.932 | (0.880,0.987) | 0.02 | 0.946 | (0.912,0.981) | **0.0029** |
| **NEU.LYM** | 0.993 | (0.964,1.023) | 0.64 | 0.990 | (0.938,1.045) | 0.72 | 0.991 | (0.956,1.027) | 0.61 |
| **MON.LYM** | 1.012 | (0.982,1.043) | 0.44 | 1.017 | (0.961,1.077) | 0.56 | 1.009 | (0.974,1.046) | 0.62 |
| **WBC.MPV** | 1.025 | (0.995,1.056) | 0.11 | 1.003 | (0.948,1.062) | 0.91 | 1.033 | (0.997,1.070) | 0.08 |
| **MPV** | 0.962 | (0.934,0.991) | **0.0096** | 0.928 | (0.877,0.983) | 0.01 | 0.975 | (0.942,1.009) | 0.15 |
| **PLT** | 1.070 | (1.040,1.101) | **3.23E-6** | 1.081 | (1.022,1.144) | 0.0062 | 1.064 | (1.030,1.100) | **2.21E-4** |
| **LYM** | 1.000 | (0.969,1.033) | 0.98 | 0.980 | (0.912,1.053) | 0.58 | 1.011 | (0.975,1.047) | 0.56 |
| **MON** | 1.005 | (0.976,1.034) | 0.75 | 0.986 | (0.930,1.045) | 0.64 | 1.011 | (0.982,1.042) | 0.45 |
| **NEU** | 0.999 | (0.970,1.029) | 0.95 | 0.975 | (0.921,1.032) | 0.39 | 1.006 | (0.971,1.043) | 0.73 |
| **HGB** | 0.922 | (0.889,0.956) | **1.03E-5** | 0.953 | (0.892,1.019) | 0.16 | 0.910 | (0.871,0.951) | **2.56E-5** |
| **RDW** | 1.010 | (0.983,1.038) | 0.46 | 1.013 | (0.953,1.076) | 0.68 | 1.008 | (0.977,1.039) | 0.63 |
| **WBC** | 1.001 | (0.970,1.032) | 0.95 | 0.969 | (0.911,1.030) | 0.31 | 1.013 | (0.977,1.051) | 0.48 |

eTable 7: Full (age, sex, BMI, smoking, and alcohol use adjusted) UKBB incident analysis overall and sex-stratified results. Boldfaced p-values indicate signals that passed FDR 0.05 threshold.

| **Trait** | **EUR N=430795** | | | **AFR N=8117** | | | **AMR N=896** | | | **SAS N=8525** | | | **EAS N=2379** | | |
| --- | --- | --- | --- | --- | --- | --- | --- | --- | --- | --- | --- | --- | --- | --- | --- |
|  | **OR** | **OR.95ci** | **p** | **OR** | **OR.95ci** | **p** | **OR** | **OR.95ci** | **p** | **OR** | **OR.95ci** | **p** | **OR** | **OR.95ci** | **p** |
| **MPV.PLT** | 0.937 | (0.909,0.967) | **4.70E-5** | 0.936 | (0.783,1.119) | 0.47 | 0.925 | (0.595,1.437) | 0.73 | 0.918 | (0.757,1.113) | 0.38 | 0.701 | (0.408,1.205) | 0.20 |
| **PLT.LYM** | 1.029 | (0.997,1.062) | 0.07 | 1.080 | (0.902,1.293) | 0.40 | 0.983 | (0.631,1.532) | 0.94 | 1.091 | (0.896,1.328) | 0.39 | 1.109 | (0.672,1.832) | 0.69 |
| **PLT.MON** | 1.011 | (0.979,1.044) | 0.49 | 1.038 | (0.871,1.236) | 0.68 | 1.443 | (1.025,2.033) | 0.04 | 1.043 | (0.859,1.266) | 0.67 | 0.733 | (0.443,1.213) | 0.23 |
| **PLT.NEU** | 1.031 | (1.000,1.064) | 0.05 | 1.069 | (0.903,1.265) | 0.44 | 1.028 | (0.691,1.529) | 0.89 | 1.233 | (1.007,1.511) | 0.04 | 1.190 | (0.717,1.973) | 0.50 |
| **HGB.RDW** | 0.949 | (0.920,0.980) | **0.0012** | 0.927 | (0.811,1.059) | 0.26 | 0.870 | (0.629,1.204) | 0.40 | 1.088 | (0.918,1.289) | 0.33 | 1.037 | (0.662,1.625) | 0.87 |
| **RDW.PLT** | 0.942 | (0.912,0.972) | **2.31E-4** | 0.870 | (0.723,1.047) | 0.14 | 0.832 | (0.515,1.343) | 0.45 | 0.973 | (0.802,1.181) | 0.78 | 0.717 | (0.413,1.245) | 0.24 |
| **NEU.LYM** | 0.998 | (0.968,1.030) | 0.91 | 1.001 | (0.841,1.190) | 0.99 | 0.961 | (0.634,1.456) | 0.85 | 0.903 | (0.733,1.112) | 0.34 | 0.942 | (0.569,1.557) | 0.81 |
| **MON.LYM** | 1.015 | (0.983,1.048) | 0.36 | 1.027 | (0.861,1.224) | 0.77 | 0.661 | (0.461,0.947) | 0.02 | 1.034 | (0.845,1.266) | 0.74 | 1.548 | (0.922,2.597) | 0.10 |
| **WBC.MPV** | 1.032 | (1.000,1.065) | 0.05 | 0.996 | (0.834,1.189) | 0.96 | 0.979 | (0.623,1.538) | 0.93 | 0.872 | (0.707,1.077) | 0.20 | 1.280 | (0.761,2.153) | 0.35 |
| **MPV** | 0.959 | (0.930,0.988) | **0.0066** | 1.132 | (0.957,1.338) | 0.15 | 1.242 | (0.848,1.820) | 0.27 | 0.773 | (0.630,0.948) | 0.01 | 0.741 | (0.425,1.294) | 0.29 |
| **PLT** | 1.069 | (1.038,1.101) | **9.08E-6** | 1.122 | (0.944,1.332) | 0.19 | 1.301 | (0.882,1.919) | 0.19 | 1.047 | (0.873,1.256) | 0.62 | 1.362 | (0.841,2.204) | 0.21 |
| **LYM** | 1.001 | (0.969,1.034) | 0.97 | 1.049 | (0.770,1.429) | 0.76 | 1.381 | (0.635,3.002) | 0.41 | 0.796 | (0.573,1.108) | 0.18 | 1.163 | (0.480,2.819) | 0.74 |
| **MON** | 1.007 | (0.979,1.036) | 0.65 | 1.060 | (0.870,1.291) | 0.56 | 0.466 | (0.217,1.000) | 0.05 | 0.876 | (0.667,1.151) | 0.34 | 1.864 | (0.949,3.661) | 0.07 |
| **NEU** | 1.005 | (0.975,1.037) | 0.73 | 1.015 | (0.826,1.247) | 0.89 | 1.174 | (0.746,1.847) | 0.49 | 0.742 | (0.586,0.940) | 0.01 | 0.994 | (0.569,1.737) | 0.98 |
| **HGB** | 0.917 | (0.883,0.953) | **8.55E-6** | 0.869 | (0.709,1.064) | 0.17 | 0.833 | (0.515,1.347) | 0.46 | 1.112 | (0.887,1.393) | 0.36 | 1.078 | (0.596,1.952) | 0.80 |
| **RDW** | 1.011 | (0.982,1.041) | 0.45 | 1.020 | (0.913,1.140) | 0.72 | 1.131 | (0.834,1.534) | 0.43 | 0.935 | (0.805,1.086) | 0.38 | 0.974 | (0.661,1.437) | 0.90 |
| **WBC** | 1.006 | (0.975,1.037) | 0.72 | 1.061 | (0.846,1.330) | 0.61 | 1.189 | (0.701,2.017) | 0.52 | 0.703 | (0.544,0.908) | 0.01 | 1.150 | (0.618,2.141) | 0.66 |

eTable 8: Full (age, sex, BMI, smoking, and alcohol use adjusted) UKBB incident analysis ancestry-stratified results. Boldfaced p-values indicate signals that passed FDR 0.05 threshold.

| Trait | OR | OR.95ci | p | N |
| --- | --- | --- | --- | --- |
| MPV.PLT | 0.971 | (0.909,1.038) | 0.39 | 452919 |
| PLT.LYM | 1.051 | (0.985,1.121) | 0.13 | 452919 |
| PLT.MON | 1.036 | (0.968,1.109) | 0.30 | 452919 |
| PLT.NEU | 1.049 | (0.984,1.119) | 0.15 | 452919 |
| HGB.RDW | 1.014 | (0.944,1.090) | 0.70 | 452919 |
| RDW.PLT | 0.988 | (0.924,1.057) | 0.73 | 452919 |
| NEU.LYM | 1.004 | (0.941,1.070) | 0.91 | 452919 |
| MON.LYM | 1.015 | (0.949,1.085) | 0.66 | 452919 |
| WBC.MPV | 0.962 | (0.902,1.026) | 0.24 | 452919 |
| MPV | 0.948 | (0.887,1.013) | 0.12 | 452919 |
| PLT | 1.012 | (0.948,1.080) | 0.72 | 452919 |
| LYM | 0.957 | (0.878,1.043) | 0.31 | 452919 |
| MON | 0.970 | (0.909,1.035) | 0.35 | 452919 |
| NEU | 0.961 | (0.901,1.026) | 0.23 | 452919 |
| HGB | 1.041 | (0.962,1.127) | 0.31 | 452919 |
| RDW | 0.999 | (0.934,1.069) | 0.98 | 452919 |
| WBC | 0.946 | (0.883,1.013) | 0.11 | 452919 |

eTable 9: Full (age, sex, BMI, smoking, and alcohol use adjusted) UKBB incident analysis sex interaction model

| **Trait** | **Overall N=3383** | | | **Male N=1643** | | | **Female N=1740** | | |
| --- | --- | --- | --- | --- | --- | --- | --- | --- | --- |
|  | **OR** | **OR.95ci** | **p** | **OR** | **OR.95ci** | **p** | **OR** | **OR.95ci** | **p** |
| **MPV.PLT** | 0.932 | (0.805,1.079) | 0.34 | 1.048 | (0.798,1.376) | 0.74 | 0.883 | (0.742,1.051) | 0.16 |
| **PLT.LYM** | 1.004 | (0.870,1.159) | 0.96 | 0.940 | (0.721,1.225) | 0.65 | 1.035 | (0.871,1.229) | 0.70 |
| **PLT.MON** | 0.951 | (0.816,1.109) | 0.52 | 0.874 | (0.651,1.172) | 0.37 | 0.995 | (0.831,1.192) | 0.96 |
| **PLT.NEU** | 1.090 | (0.943,1.259) | 0.24 | 0.945 | (0.703,1.269) | 0.71 | 1.156 | (0.979,1.365) | 0.09 |
| **HGB.RDW** | 0.909 | (0.780,1.060) | 0.22 | 1.099 | (0.787,1.536) | 0.58 | 0.875 | (0.733,1.045) | 0.14 |
| **RDW.PLT** | 0.934 | (0.806,1.083) | 0.37 | 1.021 | (0.778,1.339) | 0.88 | 0.890 | (0.746,1.062) | 0.20 |
| **NEU.LYM** | 0.935 | (0.812,1.075) | 0.35 | 0.987 | (0.739,1.319) | 0.93 | 0.910 | (0.775,1.067) | 0.25 |
| **MON.LYM** | 1.051 | (0.905,1.222) | 0.51 | 1.060 | (0.795,1.413) | 0.69 | 1.038 | (0.871,1.236) | 0.68 |
| **WBC.MPV** | 1.010 | (0.871,1.172) | 0.89 | 1.025 | (0.759,1.384) | 0.87 | 0.997 | (0.840,1.183) | 0.97 |
| **MPV** | 0.972 | (0.840,1.124) | 0.70 | 1.009 | (0.769,1.324) | 0.95 | 0.952 | (0.802,1.131) | 0.58 |
| **PLT** | 1.107 | (0.963,1.273) | 0.15 | 0.957 | (0.713,1.286) | 0.77 | 1.165 | (0.992,1.369) | 0.06 |
| **LYM** | 1.048 | (0.932,1.178) | 0.44 | 1.067 | (0.793,1.435) | 0.67 | 1.045 | (0.918,1.190) | 0.50 |
| **MON** | 1.125 | (0.972,1.302) | 0.11 | 1.106 | (0.851,1.437) | 0.45 | 1.119 | (0.939,1.334) | 0.21 |
| **NEU** | 0.988 | (0.853,1.144) | 0.87 | 1.047 | (0.776,1.413) | 0.76 | 0.957 | (0.807,1.134) | 0.61 |
| **HGB** | 0.877 | (0.733,1.049) | 0.15 | 0.998 | (0.719,1.386) | 0.99 | 0.846 | (0.680,1.054) | 0.14 |
| **RDW** | 1.038 | (0.901,1.196) | 0.60 | 0.858 | (0.615,1.195) | 0.36 | 1.079 | (0.920,1.266) | 0.35 |
| **WBC** | 1.016 | (0.877,1.176) | 0.84 | 1.083 | (0.802,1.462) | 0.61 | 0.984 | (0.829,1.167) | 0.85 |

eTable 10: Full (age, sex, BMI, smoking, alcohol use, familial relatedness, and time difference between exams adjusted) FHS incident analysis overall and sex-stratified results

Supplementary Figure 1: Simple (age and sex adjusted) and full model (age, sex, BMI, smoking, and alcohol use adjusted) UKBB ancestry-stratified results

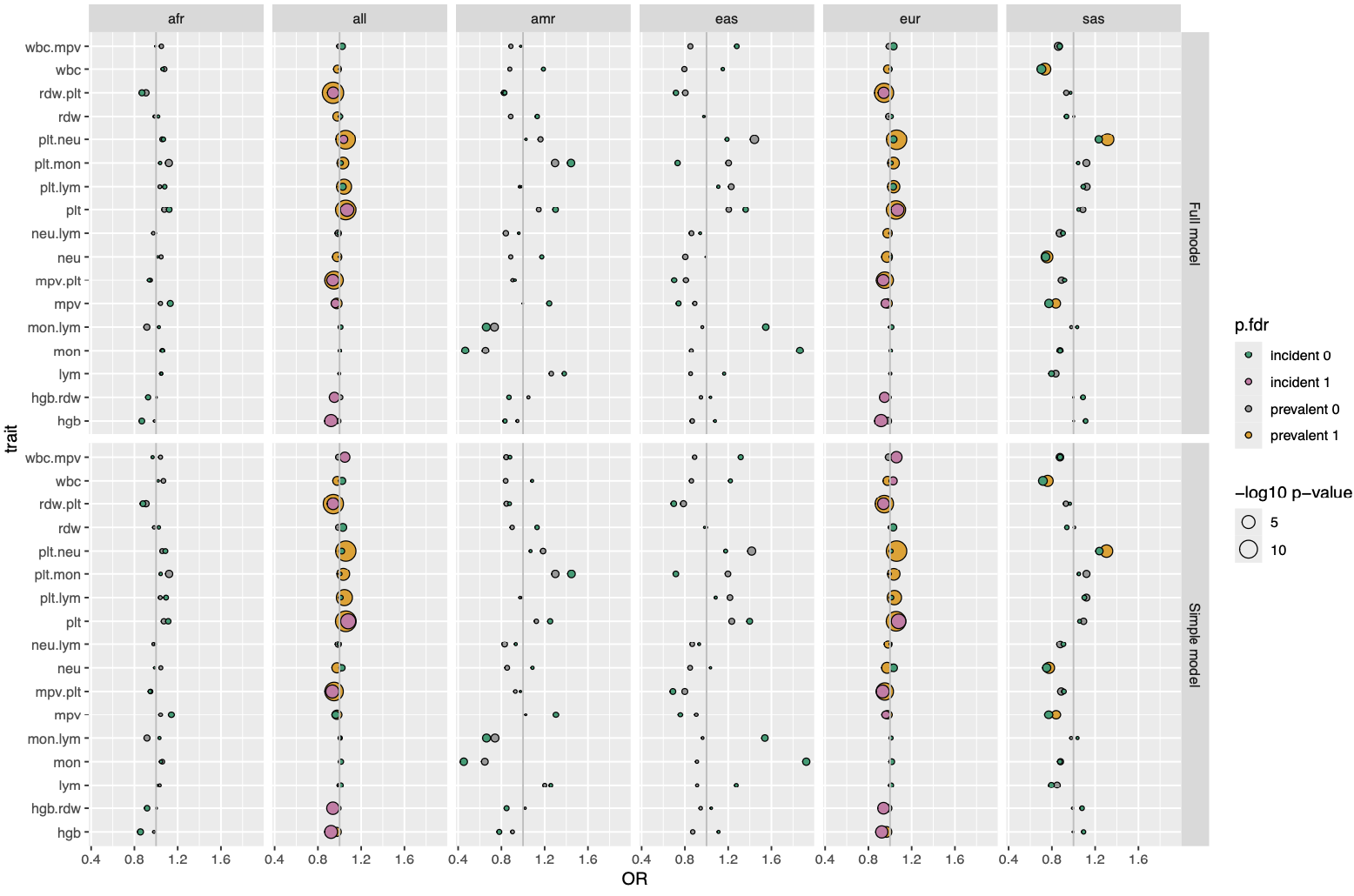
